## Supplementary table 1 for "Adipokine levels and their association with clinical disease severity in patients with dengue"

**Supplementary table 1: Clinical characteristics of dengue infection**

| **Clinical and laboratory features** | **DF (n = 49)**  **N (%)** | **DHF (n = 22)**  **N (%)** |
| --- | --- | --- |
| Fever | 49(100%) | 22(100%) |
| Headache | 38(77.5) | 19(86.36) |
| Myalgia | 30(61.22) | 22(100%) |
| Arthralgia | 24(48.9%) | 20(90.9%) |
| Nausea | 20(40.8%) | 21(95.4%) |
| Anorexia | 18(36.7%) | 20(90.9%) |
| Vomiting | 15(30.6%) | 19(86.3) |
| Pleural effusions | 0 | 0 |
| Ascites | 0 | 22(100%) |
| Bleeding | 0 | 0 |
| Shock | 0 | 0 |
| Platelet counts  <50,000  <20,000 | 13(26.5%)  4(8.1%) | 19(86.3%)  5(22.7%) |
